# Polygenic hazard score is associated with prostate cancer in multi-ethnic populations

**DOI:** 10.1101/19012237

**Authors:** Minh-Phuong Huynh-Le, Chun Chieh Fan, Roshan Karunamuni, Wesley K Thompson, Maria Elena Martinez, Rosalind A Eeles, Zsofia Kote-Jarai, Kenneth Muir, UKGPCS collaborators, Johanna Schleutker, Nora Pashayan, Jyotsna Batra, APCB (Australian Prostate Cancer BioResource), Henrik Grönberg, David E Neal, Jenny L Donovan, Freddie C Hamdy, Richard M Martin, Sune F Nielsen, Børge G Nordestgaard, Fredrik Wiklund, Catherine M Tangen, Graham G Giles, Alicja Wolk, Demetrius Albanes, Ruth C Travis, William J Blot, Wei Zheng, Maureen Sanderson, Janet L Stanford, Lorelei A Mucci, Catharine M L West, Adam S Kibel, Olivier Cussenot, Sonja I Berndt, Stella Koutros, Karina Dalsgaard Sørensen, Cezary Cybulski, Eli Marie Grindedal, Florence Menegaux, Kay-Tee Khaw, Jong Y Park, Sue A Ingles, Christiane Maier, Robert J Hamilton, Stephen N Thibodeau, Barry S Rosenstein, Yong-Jie Lu, Stephen Watya, Ana Vega, NC-LA PCaP Investigators, The IMPACT Study Steering Committee and Collaborators, Manolis Kogevinas, Kathryn L Penney, Chad Huff, Manuel R Teixeira, Luc Multigner, Robin J Leach, Lisa Cannon-Albright, Hermann Brenner, Esther M John, Radka Kaneva, Christopher J Logothetis, Susan L Neuhausen, Kim De Ruyck, Hardev Pandha, Azad Razack, Lisa F Newcomb, Canary PASS Investigators, Jay Fowke, Marija Gamulin, Nawaid Usmani, Frank Claessens, Manuela Gago-Dominguez, Paul A Townsend, William S Bush, Shiv Srivastava, Monique J Roobol, Marie-Élise Parent, Jennifer J Hu, The Profile Study Steering Committee, Ian G Mills, Ole A Andreassen, Anders M Dale, Tyler M Seibert, The PRACTICAL Consortium

**Affiliations:** Department of Radiation Medicine and Applied Sciences, University of California San Diego, La Jolla, CA, USA; Center for Multimodal Imaging and Genetics, University of California, San Diego, La Jolla, CA, USA; Division of Biostatistics and Halicioğlu Data Science Institute, University of California San Diego; Department of Family Medicine and Public Health, University of California San Diego; University of California San Diego, Moores Cancer Center, Department of Family Medicine and Public Health, University of California San Diego, La Jolla, CA 92093-0012, USA; The Institute of Cancer Research, London, SM2 5NG, UK; Royal Marsden NHS Foundation Trust, London, SW3 6JJ, UK; Division of Population Health, Health Services Research and Primary Care, University of Manchester, Oxford Road, Manchester, M13 9PL, UK; Warwick Medical School, University of Warwick, Coventry, UK; http://www.icr.ac.uk/our-research/research-divisions/division-of-genetics-and-epidemiology/oncogenetics/research-projects/ukgpcs/ukgpcs-collaborators; Institute of Biomedicine, Kiinamyllynkatu 10, FI-20014 University of Turku, Finland; Department of Medical Genetics, Genomics, Laboratory Division, Turku University Hospital, PO Box 52, 20521 Turku, Finland; University College London, Department of Applied Health Research, London, UK; Centre for Cancer Genetic Epidemiology, Department of Oncology, University of Cambridge, Strangeways Laboratory, Worts Causeway, Cambridge, CB1 8RN, UK; Department of Applied Health Research, University College London, London, WC1E 7HB, UK; Australian Prostate Cancer Research Centre-Qld, Institute of Health and Biomedical Innovation and School of Biomedical Sciences, Queensland University of Technology, Brisbane QLD 4059, Australia; Translational Research Institute, Brisbane, Queensland 4102, Australia; Australian Prostate Cancer Research Centre-Qld, Queensland University of Technology, Brisbane; Prostate Cancer Research Program, Monash University, Melbourne; Dame Roma Mitchell Cancer Centre, University of Adelaide, Adelaide; Chris O’Brien Lifehouse and The Kinghorn Cancer Centre, Sydney, Australia; Translational Research Institute, Brisbane, Queensland, Australia; Department of Medical Epidemiology and Biostatistics, Karolinska Institute, Stockholm, Sweden; Nuffield Department of Surgical Sciences, University of Oxford, Room 6603, Level 6, John Radcliffe Hospital, Headley Way, Headington, Oxford, OX3 9DU, UK; University of Cambridge, Department of Oncology, Box 279, Addenbrooke’s Hospital, Hills Road, Cambridge CB2 0QQ, UK; Cancer Research UK, Cambridge Research Institute, Li Ka Shing Centre, Cambridge UK; Population Health Sciences, Bristol Medical School, University of Bristol, BS8 2PS, UK; Nuffield Department of Surgical Sciences, University of Oxford, Oxford, OX1 2JD, UK; Faculty of Medical Science, University of Oxford, John Radcliffe Hospital, Oxford, UK; National Institute for Health Research (NIHR) Biomedical Research Centre, University of Bristol, Bristol, BS8 1TH, UK; Medical Research Council (MRC) Integrative Epidemiology Unit, University of Bristol, Bristol, BS8 2BN, UK; Faculty of Health and Medical Sciences, University of Copenhagen, 2200 Copenhagen, Denmark; Department of Clinical Biochemistry, Herlev and Gentofte Hospital, Copenhagen University Hospital, Herlev, 2200 Copenhagen, Denmark; Department of Medical Epidemiology and Biostatistics, Karolinska Institute, SE-171 77 Stockholm, Sweden; SWOG Statistical Center, Fred Hutchinson Cancer Research Center, Seattle, WA, USA; Cancer Epidemiology Division, Cancer Council Victoria, 615 St Kilda Road, Melbourne, VIC 3004, Australia; Centre for Epidemiology and Biostatistics, Melbourne School of Population and Global Health, The University of Melbourne, Grattan Street, Parkville, VIC 3010, Australia; Precision Medicine, School of Clinical Sciences at Monash Health, Monash University, Clayton, Victoria 3168, Australia; Division of Nutritional Epidemiology, Institute of Environmental Medicine, Karolinska Institutet, SE-171 77 Stockholm, Sweden; Department of Surgical Sciences, Uppsala University, 75185 Uppsala, Sweden; Division of Cancer Epidemiology and Genetics, National Cancer Institute, NIH, Bethesda, MD 20892, USA; Cancer Epidemiology Unit, Nuffield Department of Population Health, University of Oxford, Oxford, OX3 7LF, UK; Division of Epidemiology, Department of Medicine, Vanderbilt University Medical Center, 2525 West End Avenue, Suite 600, Nashville, TN 37232 USA; International Epidemiology Institute, Rockville, MD 20850, USA; Division of Epidemiology, Department of Medicine, Vanderbilt University Medical Center, 2525 West End Avenue, Suite 800, Nashville, TN 37232 USA; Department of Family and Community Medicine, Meharry Medical College, 1005 Dr. DB Todd Jr. Blvd., Nashville, TN 37208 USA; Division of Public Health Sciences, Fred Hutchinson Cancer Research Center, Seattle, Washington, 98109-1024, USA; Department of Epidemiology, School of Public Health, University of Washington, Seattle, Washington 98195, USA; Department of Epidemiology, Harvard T. H. Chan School of Public Health, Boston, MA 02115, USA; Division of Cancer Sciences, University of Manchester, Manchester Academic Health Science Centre, Radiotherapy Related Research, The Christie Hospital NHS Foundation Trust, Manchester, M13 9PL UK; Division of Urologic Surgery, Brigham and Womens Hospital, 75 Francis Street, Boston, MA 02115, USA; Sorbonne Universite, GRC n°5, AP-HP, Tenon Hospital, 4 rue de la Chine, F-75020 Paris, France; CeRePP, Tenon Hospital, Paris, France; Division of Cancer Epidemiology and Genetics, National Cancer Institute, NIH, Bethesda, MD, USA; Department of Molecular Medicine, Aarhus University Hospital, Palle Juul-Jensen Boulevard 99, 8200 Aarhus N, Denmark; Department of Clinical Medicine, Aarhus University, DK-8200 Aarhus N; International Hereditary Cancer Center, Department of Genetics and Pathology, Pomeranian Medical University, Szczecin, Poland; Department of Medical Genetics, Oslo University Hospital, 0424 Oslo, Norway; Cancer & Environment Group, Center for Research in Epidemiology and Population Health (CESP), INSERM, University Paris-Sud, University Paris-Saclay, 94807 Villejuif Cédex, France; Paris-Sud University, UMRS 1018, 94807 Villejuif Cedex, France; Clinical Gerontology Unit, University of Cambridge, Cambridge, CB2 2QQ, UK; Department of Cancer Epidemiology, Moffitt Cancer Center, 12902 Magnolia Drive, Tampa, FL 33612, USA; Department of Preventive Medicine, Keck School of Medicine, University of Southern California/Norris Comprehensive Cancer Center, Los Angeles, California, USA; Humangenetik Tuebingen, Paul-Ehrlich-Str 23, D-72076 Tuebingen, Germany; Dept. of Surgical Oncology, Princess Margaret Cancer Centre, Toronto ON M5G 2M9, Canada; Dept. of Surgery (Urology), University of Toronto, Canada; Department of Laboratory Medicine and Pathology, Mayo Clinic, Rochester, MN 55905, USA; Department of Radiation Oncology and Department of Genetics and Genomic Sciences, Box 1236, Icahn School of Medicine at Mount Sinai, One Gustave L. Levy Place, New York, NY 10029, USA; Department of Genetics and Genomic Sciences, Icahn School of Medicine at Mount Sinai, New York, NY 10029-5674, USA; Centre for Molecular Oncology, Barts Cancer Institute, Queen Mary University of London, John Vane Science Centre, Charterhouse Square, London, EC1M 6BQ, UK; Uro Care, Kampala, Uganda; Fundación Pública Galega Medicina Xenómica, Santiago De Compostela, 15706, Spain; Instituto de InvestigaciónSanitaria de Santiago de Compostela, Santiago De Compostela, 15706, Spain; Centro de Investigaciónen Red de Enfermedades Raras (CIBERER), Spain; Lineberger Comprehensive Cancer Center, University of North Carolina at Chapel Hill, Chapel Hill, NC, USA; Lineberger Comprehensive Cancer Center, University of North Carolina at Chapel Hill, 450 West Drive, CB 7295, Chapel Hill, NC 27599, USA; Department of Epidemiology, University of North Carolina at Chapel Hill, Chapel Hill, NC, USA; http://impact.icr.ac.uk; ISGlobal, Barcelona, Spain; IMIM (Hospital del Mar Medical Research Institute), Barcelona, Spain; Universitat Pompeu Fabra (UPF), Barcelona, Spain; Channing Division of Network Medicine, Department of Medicine, Brigham and Women’s Hospital/Harvard Medical School, Boston, MA 02184, USA; The University of Texas M. D. Anderson Cancer Center, 1515 Holcombe Blvd., Houston, TX 77030, USA; Department of Genetics, Portuguese Oncology Institute of Porto (IPO-Porto), Porto, Portugal; Biomedical Sciences Institute (ICBAS), University of Porto, Porto, Portugal; Univ Rennes, Inserm, EHESP, Irset (Institut de recherche en santé, environnement et travail) - UMR_S 1085, Rennes, France; Department of Urology, Mays Cancer Center, University of Texas Health Science Center at San Antonio, San Antonio Texas; Division of Epidemiology, Department of Internal Medicine, University of Utah School of Medicine, Salt Lake City, Utah, USA; George E. Wahlen Department of Veterans Affairs Medical Center, Salt Lake City, Utah 84148, USA; Division of Clinical Epidemiology and Aging Research, German Cancer Research Center (DKFZ), D-69120, Heidelberg, Germany; German Cancer Consortium (DKTK), German Cancer Research Center (DKFZ), D-69120 Heidelberg, Germany; Division of Preventive Oncology, German Cancer Research Center (DKFZ) and National Center for Tumor Diseases (NCT), Im Neuenheimer Feld 460 69120 Heidelberg, Germany; Department of Medicine, Division of Oncology, Stanford Cancer Institute, Stanford University School of Medicine, Stanford, 780 Welch Road, CJ250C, CA 94304-5769; Molecular Medicine Center, Department of Medical Chemistry and Biochemistry, Medical University of Sofia, Sofia, 2 Zdrave Str., 1431 Sofia, Bulgaria; The University of Texas M. D. Anderson Cancer Center, Department of Genitourinary Medical Oncology, 1515 Holcombe Blvd., Houston, TX 77030, USA; Department of Population Sciences, Beckman Research Institute of the City of Hope, 1500 East Duarte Road, Duarte, CA 91010, 626-256-HOPE (4673); Ghent University, Faculty of Medicine and Health Sciences, Basic Medical Sciences, Proeftuinstraat 86, B-9000 Gent; The University of Surrey, Guildford, Surrey, GU2 7XH; Department of Surgery, Faculty of Medicine, University of Malaya, 50603 Kuala Lumpur, Malaysia; Department of Urology, University of Washington, 1959 NE Pacific Street, Box 356510, Seattle, WA 98195, USA; Department of Medicine and Urologic Surgery, Vanderbilt University Medical Center, 1211 Medical Center Drive, Nashville, TN 37232, USA; Division of Medical Oncology, Urogenital Unit, Department of Oncology, University Hospital Centre Zagreb, University of Zagreb, School of Medicine, 10 000 Zagreb, Croatia; Department of Oncology, Cross Cancer Institute, University of Alberta, 11560 University Avenue, Edmonton, Alberta, Canada T6G 1Z2; Division of Radiation Oncology, Cross Cancer Institute, 11560 University Avenue, Edmonton, Alberta, Canada T6G 1Z2; Molecular Endocrinology Laboratory, Department of Cellular and Molecular Medicine, KU Leuven, BE-3000, Belgium; Genomic Medicine Group, Galician Foundation of Genomic Medicine, Instituto de Investigacion Sanitaria de Santiago de Compostela (IDIS), Complejo Hospitalario Universitario de Santiago, Servicio Galego de Saúde, SERGAS, 15706, Santiago de Compostela, Spain; University of California San Diego, Moores Cancer Center, La Jolla, CA 92037, USA; Division of Cancer Sciences, Manchester Cancer Research Centre, Faculty of Biology, Medicine and Health, Manchester Academic Health Science Centre, NIHR Manchester Biomedical Research Centre, Health Innovation Manchester, University of Manchester, M13 9WL; Case Western Reserve University, Department of Population and Quantitative Health Sciences, Cleveland Institute for Computational Biology, 2103 Cornell Road, Wolstein Research Building, Suite 2527, Cleveland, OH, 44106 USA; Uniformed Services University, 4301 Jones Bridge Rd, Bethesda, MD 20814, USA; Center for Prostate Disease Research, 1530 East Jefferson Street, Rockville, MD 20852, USA; Department of Clinical Chemistry, Erasmus University Medical Center, 3015 CE Rotterdam, The Netherlands; Epidemiology and Biostatistics Unit, Centre Armand-Frappier Santé Biotechnologie, Institut national de la recherche scientifique, 531 Boul. des Prairies, Laval, QC, Canada H7V 1B7; Department of Social and Preventive Medicine, School of Public Health, University of Montreal, Montreal, QC, Canada; The University of Miami School of Medicine, Sylvester Comprehensive Cancer Center, 1120 NW 14th Street, CRB 1511, Miami, Florida 33136, USA; http://www.cancerresearchuk.org/about-cancer/find-a-clinical-trial/a-study-find-out-lookinggene-changes-would-be-useful-in-screening-for-prostate-cancer-profile-pilot; Nuffield Department of Surgical Sciences, University of Oxford, Oxford, UK; NORMENT, KG Jebsen Centre, Oslo University Hospital and University of Oslo, Oslo, Norway; Department of Radiology, University of California San Diego, La Jolla, CA, USA; Department of Bioengineering, University of California San Diego, La Jolla, CA, USA

**Keywords:** prostate cancer, ethnicity, genetics, precision medicine, survival, hazard, risk factors

## Abstract

**Objectives:** A polygenic hazard score (PHS_1_)—weighted sum of 54 single-nucleotide polymorphism genotypes—was previously associated with age at prostate cancer (PCa) diagnosis and improved PCa screening accuracy in Europeans. Performance in more diverse populations is unknown. We evaluated PHS association with PCa in multi-ethnic populations.

**Design:** PHS_1_ was adapted for compatibility with genotype data from the OncoArray project (PHS_2_) and tested for association with age at PCa diagnosis, at aggressive PCa diagnosis, and at PCa death.

**Setting:** Multiple international institutions.

**Participants:** Men with available OncoArray data from the PRACTICAL consortium who were not included in PHS_1_ development/validation.

**Main Outcomes and Measures:** PHS_2_ was tested via Cox proportional hazards models for age at PCa diagnosis, age at aggressive PCa diagnosis (any of: Gleason score ≥7, stage T3-T4, PSA≥10 ng/mL, nodal/distant metastasis), and age at PCa-specific death.

**Results:** 80,491 men of various self-reported race/ethnicities were included (30,575 controls, 49,916 PCa cases; genetic ancestry groups: 71,856 European, 6,253 African, 2,382 Asian). Median age at last follow-up was 70 years (IQR 63-76); 3,983 PCa deaths, 5,806 other deaths, 70,702 still alive. PHS_2_ had 46 polymorphisms: 24 directly genotyped and 22 acceptable proxies (r^2^ ≥0.94). PHS_2_ was associated with age at PCa diagnosis in the multi-ethnic dataset (z=54, p<10^-16^) and in each genetic ancestry group: European (z=56, p<10^-16^), Asian (z=47, *p*<10^-16^), African (z=29, *p*<10^-16^). PHS_2_ was also associated with age at aggressive PCa diagnosis in each genetic ancestry group (p<10^-16^) and with age of PCa death in the full dataset (*p*<10^-16^). Comparing the 80^th^ and 20^th^ percentiles of genetic risk, men with high PHS had hazard ratios of 5.3 [95% CI: 5.0-5.7], 5.9 [5.5-6.3], and 5.7 [4.6-7.0] for PCa, aggressive PCa, and PCa-specific death, respectively. Within European, Asian, and African ancestries, analogous hazard ratios for PCa were 5.5 [5.2-5.9], 4.5 [3.2-6.3], and 2.5 [2.1-3.1], respectively.

**Conclusions:** PHS_2_ is strongly associated with age at PCa diagnosis in a multi-ethnic dataset. PHS_2_ stratifies men of European, Asian, and African ancestry by genetic risk for any, aggressive, and fatal PCa.

**Summary boxes:** *What is already known on this topic:*

- Genetic risk stratification can identify men with greater predisposition for developing prostate cancer, but these risk models may worsen health disparities, as most have only been validated for men of European ancestry
- A polygenic hazard score was previously associated with age at prostate cancer diagnosis and improved PCa screening accuracy in Europeans
- Performance of the polygenic hazard score in multi-ethnic populations is unknown

*What this study adds:*

- In a dataset from 80,491 men of various self-reported race/ethnicities, the polygenic hazard score was associated with age at prostate cancer diagnosis, aggressive prostate cancer diagnosis, and prostate cancer death.
- PHS stratifies men of European, Asian, and African ancestry by genetic risk for any, aggressive, and fatal prostate cancer.

## Introduction

Prostate cancer (PCa) is the second most common cancer diagnosed in men worldwide, causing substantial morbidity and mortality^1^. PCa screening may reduce morbidity and mortality^2–5^, but to avoid overdiagnosis and overtreatment of indolent disease^6–9^, it should be targeted and personalized. PCa age at diagnosis is important for clinical decisions regarding if/when to initiate screening for an individual^10,11^. Survival is another key cancer endpoint recommended for risk models^12^.

Genetic risk stratification is promising for identifying individuals with greater predisposition for developing cancer^13–16^, including PCa^17^. Polygenic models use common variants—identified in genome-wide association studies—whose combined effects can assess overall risk of disease development^18,19^. Recently, a polygenic hazard score (PHS) was developed as a weighted sum of 54 single-nucleotide polymorphisms (SNPs) that models a man’s genetic predisposition for developing PCa^13^. Validation testing was done using ProtecT trial data^2^ and demonstrated the PHS to be associated with age at PCa diagnosis, including aggressive PCa^13^. However, the development and validation datasets were limited to men of European ancestry. While genetic risk models might be important clinical tools for prognostication and risk stratification, using them may worsen health disparities^20–24^ because most models are constructed using European data and may underrepresent genetic variants important in persons of non-European ancestry^20–24^. Indeed, this is particularly concerning in PCa, as race/ethnicity is an important PCa risk factor; diagnostic, treatment, and outcomes disparities continue to exist between different races/ethnicities^25,26^.

Here, we assessed PHS performance in a multi-ethnic dataset that includes individuals of European, African, and Asian genetic ancestry. This dataset also includes long-term follow-up information, affording an opportunity to evaluate PHS for association with fatal PCa.

## Methods

### Participants

We obtained data from the OncoArray project^27^ that had undergone quality control steps described previously^18^. This dataset includes 91,480 men with genotype and phenotype data from 64 studies (**Supplemental Methods**). Individuals whose data were used in the prior development or validation of the original PHS model (PHS_1_) were excluded (n=10,989)^13^, leaving 80,491 in the independent dataset used here. **Table 1** describes available data. Individuals not meeting the endpoint for each analysis were censored at age of last follow-up.

**Table 1.** Participant characteristics, n=80,491.

|  | **Genetic Ancestry** |  |  |  |
| --- | --- | --- | --- | --- |
|  | **All** | **European** | **Asian** | **African** |
| <u>**Participants**</u> |  |  |  |  |
| Controls | 30,575 | 26,377 | 1,185 | 3,013 |
| Prostate cancer cases | 49,916 | 45,479 | 1,197 | 3,240 |
| Aggressive prostate cancer cases<sup>a</sup> | 26,419 | 24,279 | 716 | 1,424 |
| Fatal prostate cancer cases | 3,983 | 3,908 | 57 | 18 |
| <u>**Number of Participants with Known First-Degree Family History Information**</u> |  |  |  |  |
| Family history of prostate cancer available (prostate cancer cases; controls) | 46,030<br>(28,204; 17,826) | 39,445<br>(24,921; 14,524) | 1,028<br>(519; 509) | 5,557<br>(2,764; 2,793) |
| <u>**Age Demographics**</u> |  |  |  |  |
| Median age at diagnosis (IQR) | 65 [60-71] | 66 [60-71] | 68 [62-74] | 62 [56-68] |
| Median age at last follow up (IQR<sup>b</sup>) | 70 [63-76] | 70 [64-77] | 70 [63-76] | 62 [56-68] |

<sup>a</sup> Aggressive prostate cancer defined as Gleason score $\geq 7$ , PSA $\geq 10$ ng/mL, T3-T4 stage, nodal metastases, or distant metastases.
<sup>b</sup> IQR: interquartile range

All contributing studies were approved by the relevant ethics committees; written informed consent was acquired from the study participants^28^. The present analyses used de-identified data from the PRACTICAL consortium.

### Polygenic Hazard Score (PHS)

The original PHS_1_ was validated for association with age at PCa diagnosis in men of European ancestry, using a survival analysis^13^. To ensure the score was not simply identifying men at risk of indolent disease, PHS_1_ was also validated for association with age at aggressive PCa (defined as intermediate-risk disease, or above^6^) diagnosis^13^. PHS_1_ was calculated as the vector product of a patient’s genotype (*X_i_*) for *n* selected SNPs and the corresponding parameter estimates (*β_i_*) from a Cox proportional hazards regression:

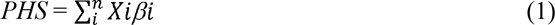

The 54 SNPs in PHS_1_ were selected using PRACTICAL consortium data (n=31,747 men) genotyped with a custom array (iCOGS, Illumina, San Diego, CA)^13^.

### Genetic Ancestry Determination

Self-reported race/ethnicities^27,29^ included European, East Asian, African American, Hawaiian, Hispanic American, South Asian, Black African, Black Caribbean, and Other. Genetic ancestry (European, African, or Asian) for all individuals was used for the present analyses because it is objective and may be more informative than self-reported race/ethnicities^30^ (**Supplemental Methods**).

### Adapting the PHS to OncoArray

Genotyping for the present study was performed using a commercially-available, cancer-specific array (OncoArray, Illumina, San Diego, CA)^18^. Twenty-four of the 54 SNPs in PHS_1_ were directly genotyped on OncoArray. We identified proxy SNPs for those not directly genotyped and re-calculated the SNP weights in the same dataset used for the original development of PHS_1_^13^ (**Supplemental Methods**).

The performance of this new, adapted PHS (PHS_2_), was compared to that of PHS_1_ in the ProtecT dataset originally used to validate PHS_1_ (n=6,411). PHS_2_ was calculated for all patients in the ProtecT validation set and was tested as the sole predictive variable in a Cox proportional hazards regression model (*R* v.3.5.1, “survival” package^31^) for age at aggressive PCa diagnosis, the primary endpoint of that study. Performance was assessed by the metrics reported during the PHS_1_ development^13^: *z*-score and hazard ratio (HR_98/50_) for aggressive PCa between men in the highest 2% of genetic risk (≥98^th^ percentile) vs. those with average risk (30^th^-70^th^ percentile). HR 95 % confidence intervals (CIs) were determined by bootstrapping 1,000 random samples from the ProtecT dataset^32,33^, while maintaining the same number of cases and controls. PHS_2_ percentile thresholds are shown in the **Supplement**.

### Any PCa

We tested PHS_2_ for association with age at diagnosis of any PCa in the multi-ethnic dataset (n=80,491, **Table 1**).

PHS_2_ was calculated for all patients in the multi-ethnic dataset and used as the sole independent variable in Cox proportional hazards regressions for the endpoint of age at PCa diagnosis. Due to the potential for Cox proportional hazards results to be biased by a higher number of cases in our dataset than in the general population, sample-weight corrections were applied to all Cox models^13,34^ (**Supplemental Methods**). Significance was set at *α*=0.01, and p-values reported were truncated at <10^-16^, if applicable^13^.

These Cox proportional hazards regressions (with PHS_2_ as the sole independent variable and age at PCa diagnosis as the outcome) were then repeated for subsets of data, stratified by genetic ancestry: European, Asian, and African. Percentiles of genetic risk were calculated as done previously^13^, using data from the 9,728 men in the original (iCOGS) development set who were less than 70 years old and without PCa. Hazard ratios (HRs) and 95% CIs for each genetic ancestry group were calculated to make the following comparisons: HR_98/50_, men in the highest 2% of genetic risk vs. those with average risk (30^th^-70^th^ percentile); HR_80/50_, men in the highest 20% vs. those with average risk, HR_20/50_, men in the lowest 20% vs. those with average risk; and HR_80/20_, men in the highest 20% vs. lowest 20%. CIs were determined by bootstrapping 1,000 random samples from each genetic ancestry group^32,33^, while maintaining the same number of cases and controls. HRs and CIs were calculated for age at PCa diagnosis separately for each genetic ancestry group.

Given that the overall incidence of PCa in different populations varies, we performed a sensitivity analysis of the population case/control numbers, allowing the population incidence to vary from 25% to 400% of that reported in Sweden (as an example population; **Supplemental Methods**).

### Aggressive PCa

Recognizing that not all PCa is clinically significant, we also tested PHS_2_ for association with age at aggressive PCa diagnosis in the multi-ethnic dataset. For these analyses, we included cases that had known tumor stage, Gleason score, and PSA at diagnosis (n=60,617 cases, **Table 1**). Aggressive PCa cases were those that met any of the following previously defined criteria for aggressive disease^6,13^: Gleason score ≥7, PSA ≥10 ng/mL, T3-T4 stage, nodal metastases, or distant metastases (**Supplemental Methods**). As before, Cox proportional hazards models and sensitivity analysis were used to assess association.

### Fatal PCa

Using an even stricter definition of clinical significance, we then evaluated association of PHS_2_ with age at PCa death in the multi-ethnic dataset. All cases (regardless of staging completeness) and controls were included, and the endpoint was age at death due to PCa. This analysis was not stratified by genetic ancestry due to low numbers of recorded PCa deaths in the non-European datasets. Cause of death was determined by the investigators of each contributing study using cancer registries and/or medical records (**Supplemental Methods**). At last follow-up, 3,983 men had died from PCa, 5,806 had died from non-PCa causes, and 70,702 were still alive. The median age at last follow-up was 70 years (IQR 63-76). As before, Cox proportional hazards models and sensitivity analysis were used to assess association.

### PHS and Family History

Family history (presence/absence of a first-degree relative with a PCa diagnosis) was also tested for association with any, aggressive, or fatal PCa. There were 46,030 men with available PCa family history data.

Cox proportional hazards models were used to assess family history for association with any, aggressive, or fatal PCa. To evaluate the relative importance of each, a multivariable model using both family history and PHS was compared to using family history alone (log-likelihood test; *α*=0.01). HRs were calculated for each variable.

## Results

### Adaption of PHS for OncoArray

Of the 30 SNPs from PHS_1_ not directly genotyped on OncoArray, proxy SNPs were identified for 22 (linkage disequilibrium ≥0.94). Therefore, PHS_2_ included 46 SNPs, total (**Supplemental Results**). PHS_2_ association with age at aggressive PCa diagnosis in ProtecT was similar to that previously reported for PHS_1_ (*z*=22 for PHS_1_, *z*=21 for PHS_2_, each *p*<10^-16^). HR_98/50_ was 4.7 [95% CI: 3.6-6.1] for PHS_2_, compared to 4.6 [3.5-6.0] for PHS_1_.

### Any PCa

PHS_2_ was associated with age at PCa diagnosis in all three genetic ancestry groups (**Table 2**). Comparing the 80^th^ and 20^th^ percentiles of genetic risk, men with high PHS had a HR of 5.3 [5.0-5.7] for any PCa. Within each genetic ancestry group, men with high PHS had HRs of 5.5 [5.2-5.9], 4.5 [3.2-6.3], and 2.5 [2.1-3.1] for men of European, Asian, and African ancestry, respectively.

**Table 2:** Association of PHS with prostate cancer. Hazard ratios (HRs) are shown comparing men in the highest 2% of genetic risk (≥98^th^ percentile of PHS), highest 20% of genetic risk (≥80^th^ percentile), average risk (30^th^-70^th^ percentile), and lowest 20% of genetic risk (≤20^th^ percentile) across genetic ancestry.

| *Genetic ancestry* | *z (p-value)* | **Hazard ratios [95% CI] comparing percentiles of PHS<sub>2</sub>** |  |  |  |
| --- | --- | --- | --- | --- | --- |
|  |  | **HR<sub>20/50</sub>:**<br>**$\leq 20^{\text{th}}$ vs 30-70<sup>th</sup>** | **HR<sub>80/50</sub>:**<br>**$\geq 80^{\text{th}}$ vs 30-70<sup>th</sup>** | **HR<sub>98/50</sub>:**<br>**$\geq 98^{\text{th}}$ vs 30-70<sup>th</sup>** | **HR<sub>80/20</sub>:**<br>**$\geq 80^{\text{th}}$ vs $\leq 20^{\text{th}}$** |
| **All**<br>**(n=80,491)** | 54 ($p < 10^{-16}$) | 0.4<br>[0.4, 0.5] | 2.4<br>[2.3, 2.5] | 4.2<br>[4.0, 4.5] | 5.3<br>[5.0, 5.7] |
| **European**<br>**(n=71,856)** | 56 ($p < 10^{-16}$) | 0.4<br>[0.4, 0.5] | 2.4<br>[2.4, 2.5] | 4.3<br>[4.1, 4.6] | 5.5<br>[5.2, 5.9] |
| **Asian**<br>**(n=2,382)** | 47 ($p < 10^{-16}$) | 0.5<br>[0.4, 0.6] | 2.2<br>[1.8, 2.6] | 3.8<br>[2.8, 5.1] | 4.5<br>[3.2, 6.3] |
| **African**<br>**(n=6,253)** | 29 ($p < 10^{-16}$) | 0.6<br>[0.6, 0.7] | 1.6<br>[1.4, 1.8] | 2.3<br>[1.9, 2.7] | 2.5<br>[2.1, 3.1] |

### Aggressive PCa

PHS_2_ was associated with age at aggressive PCa diagnosis in all three genetic ancestry groups (**Table 3)**. Comparing the 80^th^ and 20^th^ percentiles of genetic risk, men with high PHS had a HR of 5.9 [5.5-6.3] for aggressive PCa; within each genetic ancestry group, men with high PHS had HRs of 5.6 [5.2-6.0], 5.2 [4.8-5.6], and 2.4 [2.3-2.6] for men of European, Asian, and African ancestry, respectively.

**Table 3:** Association of PHS with aggressive prostate cancer. Hazard ratios (HRs) are shown comparing men in the highest 2% of genetic risk (≥98^th^ percentile of PHS), highest 20% of genetic risk (≥80^th^ percentile), average risk (30^th^-70^th^ percentile), and lowest 20% of genetic risk (≤20^th^ percentile) across genetic ancestry.

| *Genetic ancestry* | *z (p-value)* | **Hazard ratios [95% CI] comparing percentiles of PHS<sub>2</sub>** |  |  |  |
| --- | --- | --- | --- | --- | --- |
|  |  | **HR<sub>20/50</sub>:**<br>**$\leq 20^{\text{th}}$ vs 30-70<sup>th</sup>** | **HR<sub>80/50</sub>:**<br>**$\geq 80^{\text{th}}$ vs 30-70<sup>th</sup>** | **HR<sub>98/50</sub>:**<br>**$\geq 98^{\text{th}}$ vs 30-70<sup>th</sup>** | **HR<sub>80/20</sub>:**<br>**$\geq 80^{\text{th}}$ vs $\leq 20^{\text{th}}$** |
| **All (n=58,600)** | 48 ($p < 10^{-16}$) | 0.4<br>[0.4, 0.4] | 2.5<br>[2.4, 2.6] | 4.6<br>[4.3, 4.9] | 5.9<br>[5.5, 6.3] |
| **European (n=53,608)** | 46 ($p < 10^{-16}$) | 0.5<br>[0.4, 0.5] | 2.5<br>[2.4, 2.5] | 4.4<br>[4.1, 4.7] | 5.6<br>[5.2, 6.0] |
| **Asian (n=1,806)** | 44 ($p < 10^{-16}$) | 0.5<br>[0.4, 0.5] | 2.3<br>[2.2, 2.4] | 4.1<br>[3.9, 4.4] | 5.2<br>[4.8, 5.6] |
| **African (n=3,186)** | 24 ($p < 10^{-16}$) | 0.6<br>[0.6, 0.7] | 1.6<br>[1.5, 1.6] | 2.2<br>[2.0, 2.3] | 2.4<br>[2.3, 2.6] |

### Fatal PCa

PHS_2_ was associated with age at PCa death for all men in the multi-ethnic dataset (*z*=16, *p*<10^-16^). **Table 4** shows *z*-scores and corresponding HRs for fatal PCa. Comparing the 80^th^ and 20^th^ percentiles of genetic risk, men with high PHS had a HR of 5.7 [4.6-7.0] for PCa death.

**Table 4:** Association of PHS with death from prostate cancer. Hazard ratios (HRs) are shown comparing men in the highest 2% of genetic risk (≥98^th^ percentile of PHS), highest 20% of genetic risk (≥80^th^ percentile), average risk (30^th^-70^th^ percentile), and lowest 20% of genetic risk (≤20^th^ percentile).

|  |  | **Hazard ratios [95% CI] comparing percentiles of PHS<sub>2</sub>** |  |  |  |
| --- | --- | --- | --- | --- | --- |
| *Genetic ancestry* | *z (p-value)* | **HR<sub>20/50</sub>:**<br>**$\leq 20^{\text{th}}$ vs 30-70<sup>th</sup>** | **HR<sub>80/50</sub>:**<br>**$\geq 80^{\text{th}}$ vs 30-70<sup>th</sup>** | **HR<sub>98/50</sub>:**<br>**$\geq 98^{\text{th}}$ vs 30-70<sup>th</sup>** | **HR<sub>80/20</sub>:**<br>**$\geq 80^{\text{th}}$ vs $\leq 20^{\text{th}}$** |
| **All (n=78,221)** | 16 ($p < 10^{-16}$) | 0.4<br>[0.4, 0.5] | 2.5<br>[2.2, 2.8] | 4.5<br>[3.7, 5.4] | 5.7<br>[4.6, 7.0] |

### Sensitivity Analyses

Sensitivity analyses demonstrated that large changes in assumed population incidence had minimal effect on the calculated HRs for any, aggressive, or fatal PCa (**Supplemental Results)**.

### PHS and Family History

Family history was also associated with any PCa (*z*=40, *p*<10^-16^; **Table 5**), aggressive PCa (*z*=32, *p*<10^-16^), and fatal PCa (*z*=16, *p*<10^-16^) in the multi-ethnic dataset. Among those with known family history, the combination of family history and PHS performed better than family history alone (log-likelihood *p*<10^-16^). This pattern held true when analyses were repeated on each genetic ancestry. Additional family history analyses are reported in the **Supplemental Results**.

**Table 5:** Multivariable models with both PHS and family history of prostate cancer (≥1 first-degree relative affected, binary) for association with any prostate cancer in the multi-ethnic dataset, and by genetic ancestry. This analysis is limited to individuals with known family history. Both family history and PHS were significantly associated with any prostate cancer in the combined models. Hazard ratios (HRs) for family history were calculated as the exponent of the beta from the multivariable Cox proportional hazards regression^52^. The HR for PHS in the multivariable models was estimated as the HR_80/20_ (men in the highest 20% vs. those in the lowest 20% of genetic risk by PHS_2_) in each cohort. The model with PHS performed better than family history alone (log-likelihood *p*<10^-16^).

| *Genetic Ancestry* | **Variable** | **beta** | **z-score** | **p-value** | **HR** |
| --- | --- | --- | --- | --- | --- |
| **All (n=46,030)** | PHS | 2.0 | 54 | $<10^{-16}$ | 4.5 |
| | Family History | 1.0 | 39 | $<10^{-16}$ | 2.5 |
| **European (n=39,445)** | PHS | 2.1 | 56 | $<10^{-16}$ | 4.8 |
| | Family History | 1.0 | 38 | $<10^{-16}$ | 2.5 |
| **Asian (n=1,028)** | PHS | 1.9 | 51 | $<10^{-16}$ | 4.2 |
| | Family History | 0.7 | 21 | $<10^{-16}$ | 2.1 |
| **African (n=5,557)** | PHS | 1.1 | 26 | $<10^{-16}$ | 2.2 |
| | Family History | 1.1 | 47 | $<10^{-16}$ | 3.1 |

## Discussion

These results confirm the previously reported association of PHS with age at PCa diagnosis in Europeans and show that this finding generalizes to a multi-ethnic dataset, including men of European, Asian, and African genetic ancestry. PHS is also associated with age at aggressive PCa diagnosis and at PCa death. Comparing the highest and lowest quintiles of genetic risk, men with high PHS had HRs of 5.3, 5.9, and 5.7 for any PCa, aggressive PCa, and PCa death, respectively.

We found that PHS is associated with PCa in men of European, Asian, and African genetic ancestry (and a wider range of self-reported race/ethnicities). Current PCa screening guidelines suggest possible initiation at earlier ages for men of African ancestry, given higher incidence rates and worse survival when compared to men of European ancestry^26^. Using the PHS to risk-stratify men might help with decisions regarding when to initiate PCa screening: perhaps a man with African genetic ancestry in the lowest percentiles of genetic risk by PHS could safely delay or forgo screening to decrease the possible harms associated with overdetection and overtreatment^9^, while a man in the highest risk percentiles might consider screening at an earlier age. Similar reasoning applies to men of all genetic ancestries. Risk-stratified screening should be prospectively evaluated.

PHS performance was better in those with European and Asian genetic ancestry than in those with African ancestry. For example, comparing the highest and lowest quintiles of genetic risk, men with of European and Asian genetic ancestry with high PHS had HRs for any PCa of 5.5 and 4.5 times, respectively, while the analogous HR for men of African genetic ancestry was 2.5 (similar trends were seen for aggressive PCa). This suggests PHS can differentiate men of higher and lower risk in each ancestral group, but the range of risk levels may be narrower in those of African ancestry. Possible reasons for relatively diminished performance include increased genetic diversity with less linkage disequilibrium in those of African genetic ancestry^35–37^. Known health disparities may also contribute^25^, as the availability—and timing—of PSA results may depend on healthcare access. Alarmingly, there has historically been poor representation of African populations in clinical or genomic research studies^20,21^. This pattern is reflected in the present study, where most men of African genetic ancestry were missing clinical diagnosis information used to determine disease aggressiveness. That such clinical information is less available for men of African ancestry also leaves open the possibility of systematic differences in the diagnostic workup—and therefore age of diagnosis—across different ancestry populations. Notwithstanding these caveats, the present PHS is associated with age at PCa diagnosis in men of African ancestry, possibly paving the way for more personalized screening decisions for men of African descent.

The first PHS validation study used data from ProtecT, a large PCa trial^2,13^. ProtecT’s screening design yielded biopsy results from both controls and cases with PSA ≥3 ng/mL, making it possible to demonstrate improved accuracy and efficiency of PCa screening with PSA testing. Limitations of the ProtecT analysis, though, include few recorded PCa deaths in the available data, and the exclusion of advanced cancer from that trial^2^. The present study includes long-term observation, with both early and advanced disease^18^, allowing for evaluation of PHS association with any, aggressive, and fatal PCa; we found PHS to be associated with all outcomes.

Age is critical in clinical decisions of whether men should be offered PCa screening^38–40^ and in how to treat men diagnosed with PCa^38,39^. Age may also inform prognosis^39,41^. Age at diagnosis or death is therefore of clinical interest in inferring how likely a man is to develop cancer at an age when he may benefit from treatment. One important advantage of the survival analysis used here is that it permits men without cancer at time of last follow-up to be censored, while allowing for the possibility of them developing PCa (including aggressive or fatal PCa) later on. PCa death is a hard endpoint with less uncertainty than clinical diagnosis (which may vary with screening practices and delayed medical attention). PHS may help identify men with high (or low) genetic predisposition to develop lethal PCa and could assist physicians deciding when to initiate screening.

Current guidelines suggest considering a man’s individual cancer risk factors, overall life expectancy, and medical comorbidities when deciding whether to screen^6^. The most prominent clinical risk factors used in practice are family history and race/ethnicity^6,42,43^. Combined PHS and family history performed better than either alone in this multi-ethnic dataset. This finding is consistent with a prior report that PHS adds considerable information over family history alone. The prior study did not find an association of family history with age at PCa diagnosis, perhaps because the universal screening approach of the ProtecT trial diluted the influence of family history on who is screened in typical practice^13^. In the present study, family history and PHS appear complementary in assessing PCa genetic risk. Moreover, the HRs for PHS suggest clinical relevance similar or greater to predictive tools routinely used for cancer screening (e.g., breast cancer) and for other diseases (e.g., diabetes and cardiovascular disease). HRs reported for those tools are around 1-3 for disease development or other adverse outcome^44–48^; HRs reported here for PHS (for any, aggressive, or fatal PCa) are similar or greater.

Limitations to this work include that the dataset comes from multiple, heterogeneous studies, from various populations with variable screening rates. This allowed for a large, multi-ethnic dataset that includes clinical and survival data, but comes with uncertainties avoided in the ProtecT dataset used for original validation. However, the heterogeneity would likely reduce the PHS performance, not systematically inflate the results. Second, we note that no germline SNP tool, including this PHS, has been shown to discriminate men at risk of aggressive PCa from those at risk of only indolent PCa. Third, while the genetic ancestry classifications used here may be more accurate than self-reported race/ethnicity alone^30^, possible admixed genetic ancestry within individuals was not assessed; future development will consider local ancestry. As noted above, clinical data availability was not uniform across contributing studies and was lower in men of African genetic ancestry. The PHS may not include all SNPs associated with PCa; in fact, more such SNPs have been reported since the development of the original PHS^18^, some specifically within non-European populations^49–51^. Further model optimization (possibly by incorporating additional SNPs) may improve PCa risk stratification. Future work could also evaluate the PHS performance in relation to epidemiological risk factors previously associated with PCa risk beyond those currently used in clinical practice (i.e., family history and race/ethnicity). Finally, various circumstances and disease-modifying treatments may have influenced post-diagnosis survival to unknown degree. Despite this possible source of variability in survival among men with fatal PCa, PHS was still associated with age at death, an objective and meaningful endpoint. Future development and optimization hold promise for improving upon the encouraging risk stratification achieved here in men of different genetic ancestries, particularly African.

## Conclusion

In a multi-ethnic dataset comprising men of European, Asian and African ancestry, PHS was associated with age at PCa diagnosis, as well as age at aggressive PCa diagnosis, and at death from PCa. PHS performance was relatively diminished in men of African genetic ancestry, compared to performance in men of European or Asian genetic ancestry. PHS risk-stratifies men of European, Asian and African ancestry and should be prospectively studied as a means to individualize screening strategies seeking to reduce PCa morbidity and mortality.

## Data Availability

The data used in this work were obtained from the Prostate Cancer Association Group to Investigate Cancer Associated Alterations in the Genome (PRACTICAL) consortium, and from the ProtecT study. PRACTICAL consists of a collaborative group of researchers, each of whom retains ownership of their contributed data. Members of the consortium can use pooled data via proposals that are reviewed by the Data Access Committee. Approved proposals are then sent to the principal investigators of the PRACTICAL member studies, each of whom may opt to participate or not in the specific request. Readers interested in participating in the PRACTICAL consortium and gaining access to member data may find information about application at: http://practical.icr.ac.uk. ProtecT (http://www.bristol.ac.uk/population-health-sciences/projects/protect/) study data were provided by the principal investigators for the trials who retain ownership of this data; access to these data can be requested by contacting those principal investigators and submitting a request form for approval.

